# The Specchio-COVID19 cohort study: a longitudinal follow-up of SARS-CoV-2 serosurvey participants in the canton of Geneva, Switzerland (Study protocol)

**DOI:** 10.1101/2021.07.14.21260489

**Authors:** Hélène Baysson, Francesco Pennacchio, Ania Wisniak, María-Eugenia Zaballa, Nick Pullen, Prune Collombet, Elsa Lorthe, Stéphane Joost, Jean-François Balavoine, Delphine Bachmann, Andrew S Azman, Didier Pittet, François Chappuis, Omar Kherad, Laurent Kaiser, Idris Guessous, Silvia Stringhini, on behalf of the Specchio COVID19 study group

## Abstract

**Background:** The COVID-19 pandemic has affected billions of people around the world both directly through the infection itself and indirectly through its economic, social and sanitary impact. Collecting data over time is essential for the understanding of the disease spread, the incidence of COVID19-like symptoms, the level and dynamics of immunity, as well as the long-term impact of the pandemic.

**Objective:** The objective of the study was to set up a longitudinal follow-up of adult participants of serosurveys carried out in the Canton of Geneva, Switzerland, during the COVID-19 pandemic.

**Methods:** Serosurvey participants were invited to create an account on the dedicated digital platform Specchio-COVID19 (https://www.specchio-covid19.ch/). Upon registration, an initial questionnaire assessed socio-demographic and lifestyle characteristics (including housing conditions, physical activity, diet, alcohol and tobacco consumption), anthropometry, general health, and experience related to COVID-19 (symptoms, COVID-19 test results, quarantines, hospitalizations). Weekly, participants were invited to fill in a short questionnaire with updates on self-reported COVID-19-compatible symptoms, SARS-CoV-2 infection testing and vaccination. A more detailed questionnaire about mental health, well-being, risk perception, and changes in working conditions was proposed monthly. Supplementary questionnaires were proposed at regular intervals to assess more in depth the impact of the pandemic on physical and mental health, vaccination adherence, health care consumption and changes in health behaviors. At baseline, serology testing allowed to assess the spread of SARS-CoV-2 infection among the general population and subgroups of workers. Additionally, seropositive participants and a sample of randomly selected participants were invited for serologic testing at regular intervals in order to monitor both the seropersistance of anti-SARS-CoV-2 antibodies and the seroprevalence of anti-SARS-CoV-2 antibodies in the population of the Canton of Geneva.

**Ethics and dissemination:** The study was approved by the Cantonal Research Ethics Commission of Geneva, Switzerland (CCER Project ID 2020-00881). Results will be disseminated in a variety of ways, via the Specchio-COVID19 platform, social media posts, press releases, and through regular scientific dissemination methods (open-access articles, conferences).

**Article summary:** *Strengths and limitations:*

- This is a large study with a diversified recruitment among the general population and mobilized workers. It will contribute to obtain a clearer picture of the impact of the COVID-19 pandemic, for both the general population and targeted subpopulations.
- A major strength of the study is the combined use of serological testing and questionnaires. While regular serological testing will help us to model evolution of the pandemic, self-reported data on socioeconomic characteristics, COVID-19-compatible symptoms, and general and mental health will allow us to monitor the progression of the COVID-19 pandemic as well as to thoroughly analyze its effects on several dimensions of health.
- The longitudinal component of the study will provide insight into the extent and duration of immunity, as well as the long-term impact of the pandemic and the sanitary, social and economic measures associated with it.
- The main limitation is that Specchio-COVID19 is based on self-reports with a risk of information bias. However, considering the pandemic context, participants are generally engaged to participate and to contribute to COVID-19 research. Further, at least half of the sample is based on random selection in the general population.
- The study is primarily being conducted online, which may limit the generalizability of the findings, especially for the elderly and vulnerable populations, although internet access is extensive in Switzerland. Nonetheless, participants can use paper questionnaires to contribute to the major assessments.

## Introduction

Coronavirus disease 2019 (COVID-19), caused by infection with the novel coronavirus SARS-CoV-2 (1), has impacted the lives and health of billions of people around the world. First reported in Wuhan, China, in December 2019 (2), the COVID-19 epidemic has since massively spread all over the globe, accounting for more than 131 million confirmed cases and more than 3 million deaths worldwide as of June, 2021 (3). Due to the limited treatment options and and poor understanding of the virus itself, most governments have taken drastic measures to slow the epidemic spread by implementing massive population testing, quarantining of suspected cases and close contacts of infected individuals, isolation of confirmed cases, social distancing, shutting down of public infrastructures and other lockdown measures (4).

These measures are expected to have a substantial long-term impact on the population’s well-being, general and mental health and daily life, with a sudden disruption of professional, social or familial routines, as well as on national and local economies (including unemployment rates and working conditions). Psychological distress due to major uncertainties and stress related to the pandemic, as well as social isolation, may have an impact on mental health (5). Modified access to food, reduced travel and mobility, and a more sedentary lifestyle caused by the pandemic and related public health measures including lockdown and isolation/quarantine protocols, also have a strong potential for altering health behaviors, such as diet and physical activity. These changes may exacerbate in turn health and economic inequalities linked to demographic characteristics, socio-economic status, residential area, and social or clinical vulnerability, in particular for those with chronic diseases and/or mental health problems. Further, studies have shown that persistence of COVID-19 symptoms (mainly fatigue, dyspnea, and loss of taste or smell) can result in prolonged illness referred to as “long COVID” (6,7) or Post-Acute Sequelae of COVID-19 (PASC), which might disproportionately impact vulnerable population groups. It is thus of utmost importance to monitor the potential long-term complications of SARS-CoV-2 infection (6,7) as well as the consequences of potential postponement of medical interventions and screening programs due to the pandemic.

Initial screening strategies for SARS-CoV-2 infection at the beginning of the pandemic often involved limiting tests to those with severe disease or risk factors for complications, thus failing to provide information on the full extent of the epidemic. One way to solve this issue has been to conduct population-based serosurveys (8), which provide estimates of the true proportion of the population having been exposed to the virus with or without symptoms. Only longer-term clinical and serological follow-up studies will allow us to evaluate the extent and duration of immunity, to assess risk factors for infection or re-infection, and to determine the frequency and risk factors for “long COVID” symptoms among individuals who continue to report lasting effects long after being infected (6).

## Situation in the canton of Geneva and past serosurveys

The canton of Geneva, Switzerland, reported its first confirmed COVID-19 case on February 26, 2020, and by June 22, 2021, 58’889 confirmed cases and 746 deaths had been reported (9). Since the beginning of April 2020, the Geneva University Hospitals (HUG) and University of Geneva (UNIGE) have conducted repeated cross-sectional seroprevalence surveys in representative samples of the population of the canton of Geneva (10). First, participants from the ongoing population-representative “Bus Santé” study (11) were invited to participate in the SEROCoV-POP serosurvey along with their household members aged 5 years and older. The study was carried out over a total of 12 weeks (April-June 2020) and included 8’344 participants (10). A second serosurvey, SEROCoV-WORK+ (May-September 2020), recruited 10’582 essential workers who could not telework during the confinement period associated with the first epidemic wave (12). To determine population seroprevalence during the second epidemic wave, a third serosurvey was conducted during November 2020 in the framework of Corona Immunitas, a national research program assessing the seroprevalence of anti-SARS-CoV-2 antibodies across Switzerland (13). In order to be more representative of some age groups, children and their family, as well as people over 65 years were randomly selected from population registries and invited to perform serological testing. A random sample of participants from the general population who had participated in the serosurvey of spring 2020 was also invited for a second assessment. Results showed that the prevalence of anti-SARS-CoV-2 antibodies was around 11% in Geneva in May 2020 (10) and 21% by December 2020 (14). Additional serosurveys will be conducted during the follow-up period in order to obtain updated seroprevalence estimates for the general population of the canton of Geneva.

In this context, our objective was to set up a longitudinal cohort study for the long-term monitoring of adult participants from all aforementioned serosurveys. This follow-up will take place through the Specchio-COVID19 digital platform and will monitor COVID-19-related symptoms and SARS-CoV-2 seroconversion, as well as the overall impact of the pandemic on several dimensions of health and on socio-economic factors over a period of at least two years.

## Methods/Design

Follow-up of serosurvey participants using the Specchio-COVID19 digital platform will allow for the serosurveillance of the COVID-19 epidemic in Geneva as well as the continuous and dynamic monitoring of the population’s physical and mental health through online questionnaires and through repeated serological testing. More specifically, the Specchio-COVID19 study, using a dedicated digital platform, aims to:

- Monitor the epidemic and infection propagation among the general population of the canton of Geneva and among selected subpopulations;
- Identify risk factors and potentially protective factors for infection;
- Evaluate the persistence of anti-SARS-CoV-2 antibodies over time and the probability of reinfection;
- Evaluate the overall health impact of the COVID-19 pandemic in the short and long term across a large number of dimensions, including physical and mental health, behavioural changes (diet, physical activity, tobacco and alcohol consumption), forgoing healthcare, incidence of non-communicable diseases in the population, and persistence of COVID-19-related symptoms (“long COVID”);
- Evaluate the overall socio-economic impact on the population of the COVID-19 pandemic, including in the long term;
- Evaluate risk perception, the adoption of preventive behaviors and acceptance of COVID-19-related public health policies over time;
- Collaborate as part of the Corona Immunitas national research program (https://www.corona-immunitas.ch/), coordinated by the Swiss School of Public Health (SSPH+) which aims to generate comparable seroprevalence estimates across Switzerland (13).

### Study design

In order to quickly respond to the COVID-19-related health emergency, and thus support public health actors in decision-making, we decided to take advantage of an online digital platform of the population of Geneva (Specchio) developed through a collaboration between the University of Geneva and the Health Directorate of the Canton of Geneva (15). The Specchio-COVID19 internet platform allows prospective longitudinal follow-up of participants of serosurveys in the canton, resulting in the constitution of the Specchio-COVID19 digital cohort. The Specchio-COVID19 cohort will be followed-up for at least 2 years.

### Recruitment and inclusion

Participants of the Specchio-COVID19 platform are currently being recruited among adult participants of serosurveys conducted in Geneva. Progressively, we plan to extend the recruitment to other targeted populations in Geneva. All participants over the age of 18 for whom an e-mail address is on file receive an invitation to create a personal account on the Specchio-COVID19 digital platform. Informed consent is obtained during the baseline serology testing visit.

Upon registration on the digital platform, participants are included in the study and then followed via a website specifically created for this purpose. Questionnaires can be directly filled-in online using a secure HTLM interface, where all requirements to guarantee the secured management and storage of sensitive data are met. In order to be included in the cohort, participants have to fill in a mandatory initial questionnaire. After this inclusion step, other questionnaires related to the COVID-19 pandemic and its impact on physical and mental health and wellbeing are proposed to participants on a regular basis. The content of these questionnaires is detailed below in sections 3 “Digital Follow-up” and 4 “Additional questionnaires”. Serological tests were performed on all participants at baseline and are repeated over time for subgroups of participants, selected according to the needs of the study at each stage (see section 2).

## Data collection techniques

### 1 Information collected at inclusion

At inclusion, via an initial questionnaire completed on-line by the participant, information is collected on sociodemographic characteristics, general heath, presence of chronic diseases and physical or mental disability, medication use and health behaviors (diet, physical activity, alcohol and tobacco consumption, forgoing healthcare). Regarding socio-demographic characteristics, information is collected regarding residential postal code, household composition (number of co-habitants, number of children) and housing conditions, work situation (employment status, profession), education level, and income. As the canton of Geneva is cosmopolitan with 40 percent of residents and workers of foreign origin (16), additional information is collected about nationality, ethnicity and mother tongue. Subjective health status (perceived general health and perceived mental health), current weight, vaccination status and pregnancy status are also collected at inclusion.

### 2. Biological and clinical follow-up

#### 2a. Follow-up of seropositive participants

Seropositive participants are invited for serological testing at regular intervals, regardless of their symptomatology. This allows us to monitor the persistence of anti-SARS-CoV-2 antibodies over time. A first seropersistence survey was conducted in November 2020, showing seropersistence of anti-SARS-CoV-2 antibodies in 100% of participants at 115-224 days after their baseline serological test, as measured by the Elecsys® anti-RBD assay (Roche Diagnostics, Rotkreuz, Switzerland) (16).

#### 2b. Follow-up of randomly selected participants

Depending on the evolution of the COVID-19 epidemic in Geneva and in coordination with the Corona Immunitas national research program, samples of randomly selected participants from population registries and among our longitudinal cohort participants are repeatedly invited for serological testing, regardless of their symptomatology or previous serological status, in order to monitor the seroprevalence of anti-SARS-CoV-2 antibodies in the general population over time.

#### 2c. Serological testing

Assays used for serological testing may vary during the course of the study due to new, more performant tests being commercialized. At baseline or follow-up visits of our study, SARS-CoV-2 antibodies have been measured using three commercially-available tests: a semiquantitative anti-S1 ELISA detecting IgG (Euroimmun, Lübeck, Germany #EI 2606-9601 G, referred to as EI), and the quantitative Elecsys anti-RBD (#09 289 275 190, Roche-S) and semiquantitative Elecsys anti-N (#09 203 079 190, Roche-N), both measuring IgG/A/M levels (Roche Diagnostics, Rotkreuz, Switzerland). The assessment of antibodies against the spike protein (anti-S) and the nucleocapsid protein (anti-N) of SARS-CoV-2 permits, in principle, the distintiction between individuals with past infection from those who were vaccinated against COVID-19 (as mRNA-based vaccines are the only ones so far approved in Switzerland). All serological tests are centralized and performed blinded to participants’ characteristics or clinical history.

### 3 Digital follow-up

The digital follow-up consists of brief weekly updates with respect to self-reported symptoms compatible with COVID-19, SARS-CoV-2 testing (RT-PCR or serological tests), and vaccination status. In a more detailed monthly follow-up, questions are addressed about general and mental health, well-being and risk perception, changes in health behaviors (diet, physical activity, alcohol and tobacco consumption, sleep quality), and changes in work conditions or employment. Regarding mental health, PHQ-2 (17), GAD-2 (18) and K6 (19) scores are used to assess respectively depression anxiety and psychologic distress. Questions about social isolation and feelings of loneliness are also asked.

### 4 Additional questionnaires

### 4a. General and mental health assessed respectively every 3 and 6 months

Questions about long-term COVID-19-related symptoms are addressed in order to better characterize long COVID illness. New health events (newly diagnosed chronic diseases, new treatments, hospitalizations) are also recorded.

### 4b. Family life and children

Participants who indicate having children under 18 are invited to answer additional questions about their children’s stress levels and their parental experiences during the pandemic (in line with home schooling and school closures).

### 4c. Access to health care

Specific questionnaires will assess the effects of the COVID-19 pandemic on the utilization of health and social care services, and medication adherence.

### 4d. Employment and working conditions

A specific questionnaire will be developed on employment status, working conditions and financial status to get an overview of the economic impact of the pandemic in the population of Geneva, including subgroups of essential workers who could not telework during (semi-)lockdown periods.

### 4e. Vaccination

A specific questionnaire assesses acceptability and uptake of the vaccine against SARS-CoV-2 over time.

## Participants and public involvement

Given the high frequency of questionnaires to be filled in and the long duration of the follow-up period, several measures were designed to maintain high retention of participants. These measures include regular electronic newsletters with links to the “News” Specchio-COVID19 webpage, containing short videos describing the study and novel findings, media releases and accessible summaries of scientific articles published as part of the Specchio-COVID19 project. The organization of webinars specifically dedicated to participants is also planned. These activities are specifically designed to inform the participants and motivate them to remain in the study. Regarding participant involvement, questionnaires include open comment fields in which participants can describe their experiences related to the pandemic or to the study questionnaires. A specific email address and a dedicated hotline were set up so that participants can get in touch with the Specchio-COVID19 team. Participants receive personal answers in case of technical difficulties or any questions about the study or their serological results. Participants’ comments are considered when improving revised versions of the questionnaires. Other examples of participant communication include result visualizations on the national research program Corona-Immunitas website (https://www.corona-immunitas.ch/), coordinated by the Swiss School of Public Health (SSPH+) and short videos on the Corona-Immunitas’ YouTube channel (e.g., Science in A Minute, …), which are regularly updated.

## Statistical analysis plan

We plan to use this study to conduct timely data collection and analyses meeting public health or scientific needs. In this context, new scientific questions may arise regarding the impact of the pandemic and the evolving scientific knowledge about SARS-CoV-2 transmission. When new questions will arise, we will develop analysis plans prior to collecting data through new questionnaires and biological testing and undertaking analyses. The data collected will be analyzed for the entire study population and for subgroups defined by demographics, comorbidities or clinical characteristics. Through the serosurveys conducted in the canton of Geneva and the national research program Corona Immunitas, we anticipate enrolling at least 10’000 participants. The following section provides an overview of ‘basic’ analytical strategies that will be conducted with the data. Entered data will be summarized first by using simple descriptive statistics. Then, appropriate statistical methods will be used, especially Chi-2 tests and general linear models. Bayesian regression models will be performed for estimating population seroprevalence by accounting for age, sex and test performance. Analyses will be conducted in R (R Foundation for Statistical Computing, Vienna, Austria) or STATA (StataCorp LLC, Texas, USA) environments.

## Ethical considerations

The purpose of this study is explained to all individuals at the time of their first serological testing performed in the context of one of the serosurveys, all approved by the Cantonal Research Ethics Commission of Geneva, Switzerland (CCER project ID 2020-00881 and PB_2016-00363). Informed signed consent is obtained for participation in the Specchio-COVID19 study. Personal identifying information is stored in a password-protected database containing only contact details. The data from questionnaire responses and biological results are stored in a separate password-protected anonymous SUGAR-CRM database (https://www.sugarcrm.com). Only the administrators have access to the identity of the participants, in order to organize visits for biological testing. The data is stored on secure servers in Switzerland for as long as they are needed to achieve the study objectives. Compliance with data protection regulations was approved by the Cantonal Research Ethics Commission of Geneva, as part of the SEROCoV-WORK+ study protocol (CCER Project ID 2020-00881).

## Dissemination

Due to the evolving nature of the COVID-19 pandemic, results are disseminated in a variety of ways. As mentioned earlier, results are communicated through the Specchio-COVID19 platform via dedicated webpages (“News” page, “Research” pages) and via an electronic newsletter which is regularly sent by email to participants and collaborators. Where appropriate, press releases and social media are targeted. Results are also released through traditional scientific dissemination methods, journal articles, open-access publications and conference presentations. Of note, the Specchio-COVID19 study, integrated in the Corona Immunitas national research program, also provides data to the Swiss Federal Office of Public Health and to the Swiss COVID-19 scientific task force addressing key questions concerning seroprevalence of anti-SARS-CoV-2 antibodies, population adherence to preventive measures (facemasks, social distancing) and COVID-19 vaccine uptake.

## Discussion

Thanks to the Specchio-COVID19 platform, we will continue to monitor the evolution of anti-SARS-CoV-2 seroprevalence in the population of the canton of Geneva. Longer-term clinical and serological follow-up via the Specchio-COVID19 platform will allow us to monitor the duration of anti-SARS-CoV-2 antibodies and the possible clinical sequelae of COVID-19 over time. Data collected via Specchio-COVID19 will also permit more detailed analyses on the seroprevalence distribution within the population, taking into account symptomatology and sociodemographic factors, to better understand transmission and selective risk of infection within the community. Immunity dynamics related to anti-SARS-CoV-2 antibodies are still largely unknown. For example, whether the presence of anti-SARS-CoV-2 antibodies protects individuals from a new COVID-19 infection, especially in relation to the circulation of new variants, is unknown at this stage. If it does, the duration of immunity also needs to be evaluated. The duration of seroprotection of vaccinated individuals also remains unknown at the population level. Following a large and diversified cohort, with blood samples collected and stored for serological analyses since the beginning of the pandemic, will allow us to address these questions.

Detailed information on the effects of the pandemic on mental and physical health, as well as on social and professional life are also collected. Although monitoring of COVID-19 patients and contact-tracing is assured by the official state department in charge, independently from our study, expected results will help design adequate public health policies to manage new potential outbreaks of the COVID-19 pandemic but also to face the possible clinical complications that may occur after the acute phase of the disease subsides (long COVID) and to prepare the healthcare system for such challenges. The long-term implications of SARS-CoV-2 infection on morbidity and mortality are as of yet unknown, although some basic deductions can be made based on clinical experience gained from residual long-term effects of SARS and MERS epidemics (20). Long-term complications need to be carefully followed up over time. Moreover, indirect long-term effects of public health measures on health, such as the effects of social distancing on mental health or the consequences of healthcare renunciation on chronic disease management, are also expected.

## Strengths

Strengths of our project pertain to its large sample size and recruitment of participants across different populations (general population, non-confined workers). Around 20’000 participants already included in either one of the SEROCoV-POP, SEROCoV-WORK+, and Corona Immunitas Geneva serosurveys have been invited by e-mail to join the Specchio-COVID19 platform at the end of 2020. Assuming a participation rate of 50%, we estimate that around 10’000 participants will be included by the end of 2021, leading to a cohort of participants that will be followed over time. It is also planned to carry out a specific study focusing on children (SEROCoV-KIDS), who will be integrated in to our digital cohort, and to open the recruitment more extensively through time, depending on scientific needs. The flexibility of the Specchio-COVID19 web platform allows for a quick implementation of new questionnaires to provide insights into multiple aspects of this singular situation at a large-scale level, while adapting to the evolving phases of the pandemic. Previous initiatives in Switzerland, such as Grippenet (21), have demonstrated the potential for using online surveys for disease surveillance (22). Another strength of the Specchio-COVID19 project is the collaboration of researchers across a wide range of disciplines (epidemiologists, infectious diseases specialists, medical doctors, psychologists, biologists, data scientists and statisticians) in designing and implementing questionnaires, enabling a broad spectrum of research questions to be addressed. One of the major strengths is that Specchio-COVID19 combines data from self-reported questionnaires and results from biological testing. Self-reported data on socioeconomic characteristics, symptoms, and contact tracing provides understanding of the pandemic in more depth than serological testing alone. It will enable assessment of the severity of illness more accurately, as well as transmission dynamics, and the effect of socioeconomic characteristics.

As a subsample of the participants of Specchio-COVID19 have also been included in the “Bus Santé” study (11), an annual health survey of a representative sample of the population of the canton of Geneva which has been carried out for almost 30 years, it is possible to compare data collected through paper questionnaires and face to face clinical interviews (in the “Bus Santé” study) with data collected through online questionnaires (via the Specchio-COVID19 platform). This will allow us to provide an insight into declaration bias for social desirability answers (weight, tobacco consumption, etc.) and to acquire data on behavioral changes before and after the pandemic for a subsample of our participants.

## Limitations

Some limitations should be acknowledged. Specchio-COVID19 is based on voluntary involvement. Reasons why individuals might not participate include lack of time and motivation, poor internet access, language restrictions or a poor understanding of the purpose of the study. As in previous studies with a comparable design (23), it is expected that more women than men and highly educated participants with a high socio-economic status will participate in comparison with the general population of the canton of Geneva. To increase participation rate among socially vulnerable or less educated populations, targeted recruitment will be carried out with the support of associations in Geneva. Web-based participation might exclude less technology-proficient individuals. To mitigate this limitation, it is planned to send paper questionnaires at less frequent intervals to individuals who make such a request. With time, we will provide study documents in languages other than French and provide a multi-language platform. To minimize social desirability bias (for example regarding health preventive behaviors), questionnaires are web-based and anonymity is guaranteed. Finally, we plan to use our original platform Specchio (15) after the pandemic ends to continue following participants of Specchio-COVID19 for the coming decade and beyond, providing opportunities to examine the long-term health impacts of the pandemic.

As for any other long-term prospective cohort, attrition remains a major concern. Over time, willingness to participate to the study may diminish considerably and low response rates might affect the representativeness of the study population, the generalizability of the findings, and the power of the statistical analyses. In order to favor its use and maintain participation over time, the online platform Specchio-COVID19 is designed to be attractive and user-friendly. E-mail reminders will be sent to encourage participation, and in case of non-response or incomplete questionnaires. Motivational strategies will be implemented to improve retention of participants, for example “digital awards” (e.g. a trophy sticker) or participant certificates. Communication with participants will be reinforced through regular updates on the progress and main achievements of the project *via* the Specchio-COVID19 platform and a periodic newsletter. A team specifically trained and dedicated to Specchio-COVID19 informs and responds to participants’ queries. As the research topic is of utmost interest for many people in the current situation of a pandemic, the response rate is expected to be higher in the Specchio-COVID19 study than in other non-COVID-19-related studies. However, one key issue is that participants will be aware of their serological status during follow-up, which may affect behavior and risk perception, and therefore willingness to participate in the study over time.

### Links to other national and international research programs

In order to have a broader overview of the consequences of the pandemic beyond Geneva, Specchio-COVID19 collaborates with the Corona Immunitas national research, coordinated by the Swiss School of Public Health (SSPH+). This national program aims to harmonize ongoing seroprevalence studies across Switzerland, targeting the population as a whole or particular subgroups, such as children, seniors, health professionals, essential and non-confinable workers, or vulnerable populations. Comparable results of these studies will provide an accurate view of the proportion of the Swiss population infected with the virus and of the impact of the pandemic on several dimensions of health (13). Comparisons with international projects carried out in several European countries are possible, and will provide unique opportunities to examine the effect of different governmental measures on seroprevalence, mental health and well-being of the population and on the economical and societal impact of the pandemic worldwide. In Germany, the KoCo19 (prospective Covid-19 cohort) study included more than 5000 participants drawn from a random sample of 3000 households in Munich (24). After a baseline visit conducted between April and June 2020, participants completed an online household questionnaire and an online personal questionnaire, a daily digital diary about symptoms, outings, use of public transportation, social contacts as well as additional questions about the psychosocial and economic situation. In France, the Sapris-SERO study was implemented in March 2020 to evaluate the main epidemiological, social and behavioral challenges of the SARS-CoV2 epidemic in France in relation to social inequalities in health and healthcare. It is based on a consortium of 16000 participants selected from ongoing prospective cohort studies including three general population-based adult cohorts and two child cohorts (25). In Luxembourg, 1862 participants were followed since April 2020 as part of the CON-VINCE study, a national survey of randomly selected asymptomatic adults (26).

## Conclusion

Considering its large size and the wealth of data collected through questionnaires combined with serosurveillance, the Specchio-COVID19 digital cohort constitutes a powerful tool for public health information and epidemiologic surveillance in the context of the COVID-19 pandemic. The follow-up of participants’ serological status provides data on the proportion of the population with anti-SARS-CoV-2 antibodies at different time points (combining information on the evolution of the epidemic and the progression of the vaccination campaigns) as well as on the lifetime of these antibodies (and potentially the immunity conferred by them). These data are essential for the implementation of any preventive measures aimed at managing new epidemic waves and an overload of healthcare facilities. The data collected through questionnaires provide an overview of the psychological, societal and economic impacts of the pandemic on the population in general and on subgroups which may have been particularly exposed either clinically, socially or economically. These data will also help health policy makers in developing prioritized actions during and after the pandemic.

**Figure 1.**
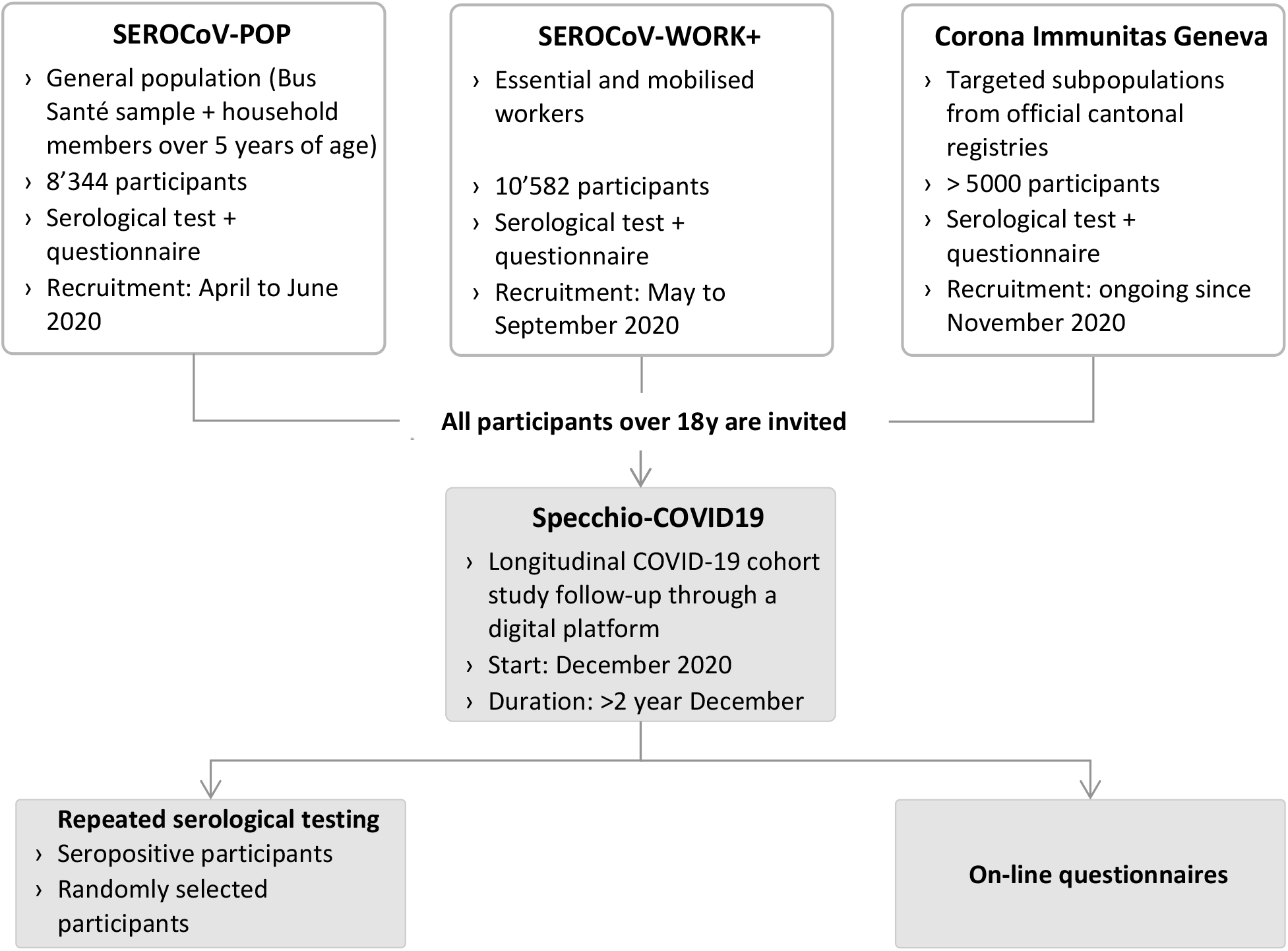
Design of Specchio-COVID19 study.

**Figure 2.**
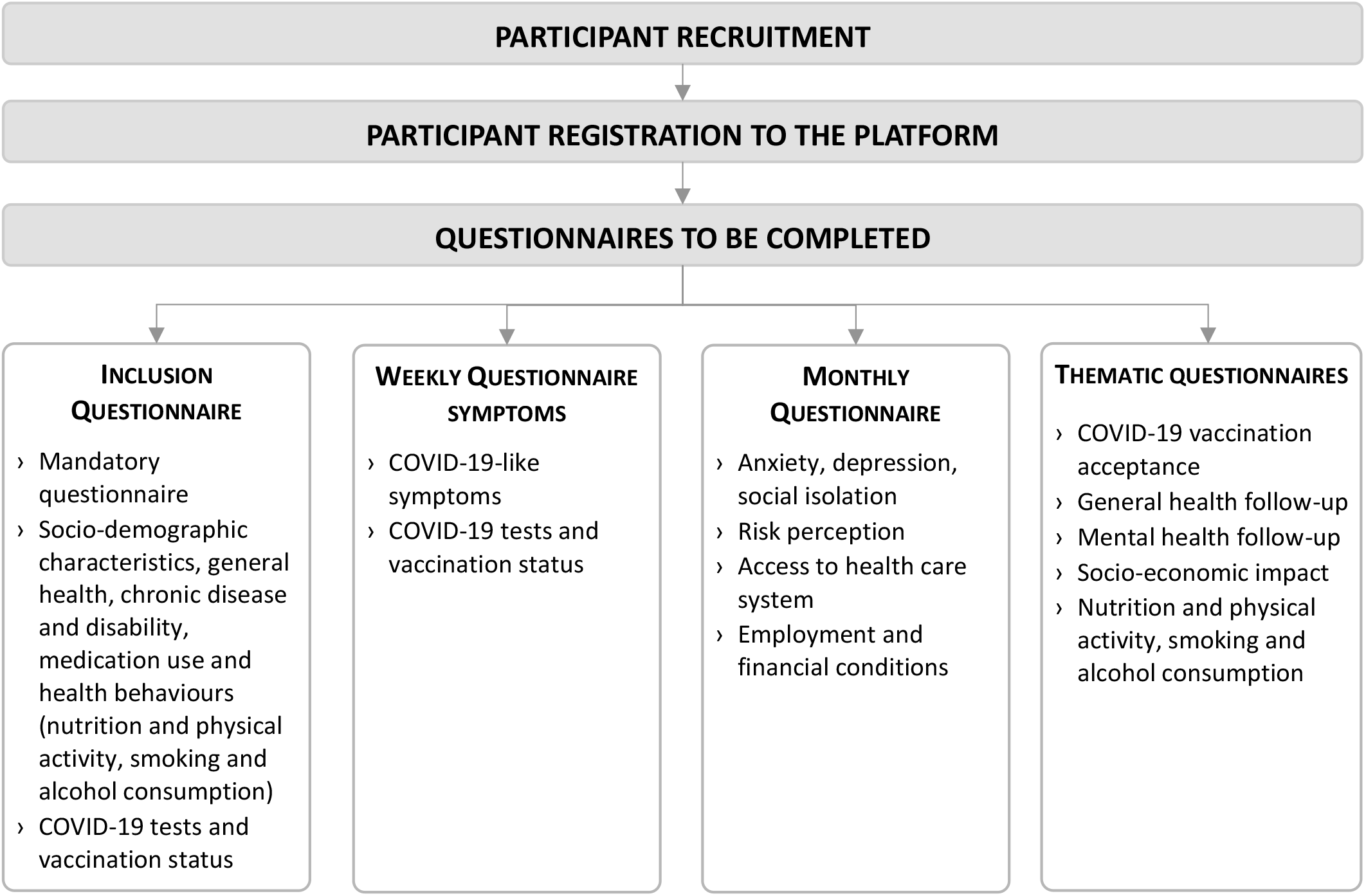
Recruitment and questionnaires.

**Figure 3.**
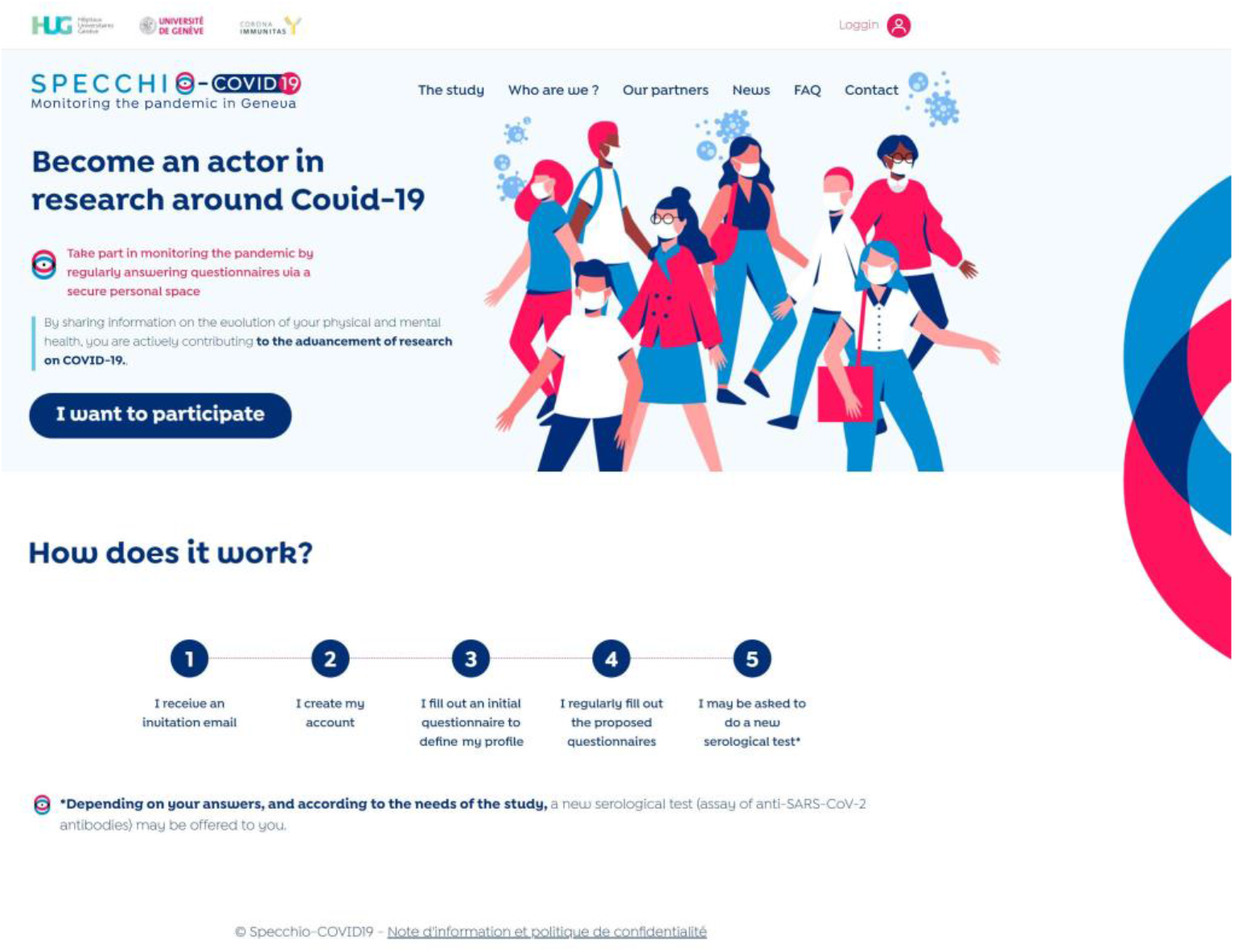
Home page of Specchio-COVID19 platform translated from French, www.specchio-COVID19.ch (Only available in French at the time of submission)

## Data Availability

Data are available to the scientific community upon submission of a data request application to the investigators board via the corresponding author.

## Acknowledgments

The authors are indebted to the participants of the Specchio-COVID19 cohort for their commitment and cooperation. This study would not have been possible without the instrumental and passionate contribution of the staff of the Unit of Population Epidemiology of the Primary Care Division, Geneva University Hospitals, Geneva, Switzerland and the professionalism of our partners Med&Hyg and DotBASE, and all our colleagues not included in the Specchio-COVID19 study group. We particularly thank the Specchio-COVID19 operational team who spend a dedicated time responding to participants’ queries.

## Authors’ contribution

All authors participated in the study design and helped to draft the final version of the manuscript, contributing substantially to conception and design of the study; drafting the article and revising it critically for important intellectual content; providing final approval of the version to be published; and acting as guarantors of the work.

All authors read and approved the final manuscript

## Conflicts of interest to disclose

The authors declare that they have no competing interests.

## Data statement

Participants’ informed consent does not authorize data, even coded, to be immediately available. It does allow, however, for the data to be made available to the scientific community upon submission of a data request application to the investigators board via the corresponding author.

